# Pharmaceutical Repurposing Strategies for Metabolic Disorders: Insights from Mendelian Randomization Studies

**DOI:** 10.1101/2024.03.29.24305070

**Authors:** Shixuan Zhang, Xiaoxi Hu, Dandan Hou, Xiaoru Sun, Dayan Sun

## Abstract

Metabolic disorders (MDs) are a group of medical conditions that impact the metabolism. These complex processes may have common characteristics among various diseases, thereby suggesting the potential of drug repurposing. Employing Mendelian Randomization (MR), we constructed a causal network between 2,478 targetable drug-gene expressions (eQTLs) and 30 broadly reported metabolic disorders. Our study identified 499 drug target genes significantly associated with 27 metabolic disorders (|MR coefficient| > 0.2). Pathway enrichment analysis of drug target genes indicates that regulation of response stimulus may serve as a common pathway across 14 diseases. Based on 53 commonly used clinical drugs for 18 diseases, we elucidated novel therapeutic mechanisms of some drugs, such as the potential of Valproic Acid to treat Schizophrenia by affecting key genes in Alcoholism, *SLC29A1*, and *HDAC4*. Furthermore, we identified potential for drug repurposing in four diseases, with Manic Episode and Type 1 Diabetes sharing four novel drugs: Cannabidiol, Doxorubicin, Genistein, and Propylthiouracil. Additionally, we predicted 189 potential therapeutic drugs affecting the causal genes of diseases. Overall, we established a causal network between metabolic disorders and drug target genes, explored possible pathways for drug action on disease treatment, and proposed drug repurposing strategies for four diseases.

**Highlights:**

1. To the best of our understanding, the manuscript we have submitted delineates the inaugural drug-gene causal network tailored for Metabolic Disorders (MDs), covering an array of 30 diseases.
2. This investigation unveils potential novel drug mechanisms aimed at treating MDs, marking a pioneering exploration in this domain.
3. We introduce strategies for the repurposing of drugs targeting four specific MDs, suggesting innovative approaches to treatment.
4. The findings are presented through an easily navigable web interface, enabling thorough examination by users (https://quantlifebits.shinyapps.io/mrmetabopharm/).

## Introduction

Metabolic disorders are emerging as a significant global health concern, with their prevalence experiencing a marked surge^1^. This growing interest is attributed to their extensive systemic complexity and widespread occurrence. The excessive variation in the human body’s internal environment is a primary trigger for these diseases, such as endocrine disruptions in hormones (diabetes^2^, hypertension^3,4^, dyslipidemia^5^, obesity^6^, Polycystic Ovary Syndrome^6^, etc.), disruptions in circadian rhythms (depression^7^, Alzheimer’s disease^8^, etc.), and dysregulation of cell growth regulation, all of which may be associated with chronic systemic diseases involving multiple organ systems.

Metabolic disorders may exhibit potential bidirectional interactions, thus offering ample opportunities for drug repurposing. Notably, diabetes is concurrently associated with several diseases in a bidirectional manner, such as cardiovascular and cerebrovascular diseases^9^, behavioral disorders^10^, cancer^11^, and dementia^12^, with diabetes medication metformin being utilized to assist in treating these conditions^13–15^. The accumulation of metabolic disorder states may lead to organ system damage, a process akin to aging^16^, affecting systems such as the cardiovascular^17^, nervous^18^, and endocrine systems^19^, with more severe accumulations potentially leading to various cancers^20^. Furthermore, Wray NR et al. suggested that the pathogenesis of metabolic diseases is complex^21^, and different diseases may share common pathogenetic mechanisms. Further research indicated that comorbidities of many diseases might be related to genetic factors^22^, suggesting the possibility of common effective intervention mechanisms between different diseases. Therefore, employing Mendelian randomization to identify common causal drug target genes may be an effective approach for developing new therapeutic strategies for diseases^23^.

The potential efficacy of classic drugs such as metformin in treating metabolic disorders other than diabetes underscores the significance of the interconnectedness among metabolic disorders. In 1988, Black J. proposed drug repurposing schemes^24,25^, which hold significant advantages in shortening the drug development cycle, mitigating disease resistance sooner^26^, and avoiding safety issues associated with new drug development^27^. While numerous treatment options exist for diseases classified as metabolic disorders ^1,28,29^, the need for further refinement of treatment strategies is underscored by the increasing prevalence rates published by the World Health Organization (WHO)^1,30^. Consequently, the broadening of existing drugs’ applications to encompass various diseases through shared genetic effects and causal genes, alongside investigating the mechanisms underlying drug actions, has become a focal point of research^31^. This approach is vital for circumventing issues such as drug resistance, adverse effects, and safety concerns associated with new medications in managing metabolic disorders. It holds significant promise for enhancing therapeutic strategies.

In this study, we explored potential drug-regulatory genes for diseases classified as metabolic disorders based on blood-derived targetable drug genes (expression Quantitative Trait Loci, eQTLs). The research design is depicted in **Fig. 1**. Initially, we utilized Genome-Wide Association Study (GWAS) data from FinnGen, eQTLs from the eQTLGen Consortium, and the Drug-Gene Interaction database to establish a complex causal network between drug genes and multiple diseases. Subsequently, methods such as reverse causation detection, pathway enrichment analysis, and drug prediction were employed to validate the primary findings. Furthermore, we constructed an effective causal network of ‘drug-gene-disease’ for metabolic disorder diseases and performed external validation for some of the diseases included in the analysis using GWAS data from the UK Biobank cohort to enhance the level of evidence. Finally, we have defined the network we constructed as “Metabolic disorders targetable drug causal network,” publicly available on an interactive website (https://quantlifebits.shinyapps.io/mrmetabopharm/) to serve as a navigational map for subsequent research.

## Methods

### Data inclusion

#### eQTL of drug gene collection

For the assembly of drug gene sets, 4,464 genes associated with known or potential drugs were selected from the Drug-Gene Interaction database. Significant SNP loci within ±100 kb of the start and end positions of each gene were extracted to form a set of genetic variations. Using peripheral blood meta-analysis cis-eQTL data (within ±1Mb of each probe and p value < 0.05 after false discovery rate correction) from 31,684 samples via the eQTLGen Consortium platform, SNPs significantly associated with drug genes within the genetic variation set (p value < 5X 10^-8^) were identified as genetic instrumental variables. The corresponding eQTL results represent summary statistical data between genetic variants and genes. SNPs with a Minor Allele Frequency (MAF) greater than 1% and an F-statistic greater than 10 (based on MAF) were retained to mitigate the bias from weak instrumental variables. These variants were then clumped with an R^2^ of 0.1% and a window size of 10,000 kb, based on the reference data from the European population in the 1000 Genomes Project. The research ultimately incorporated eQTLs from 2,478 drug-targetable genes for further investigation.

#### Disease set screening

The disease collection was selected based on keywords including metabolic disorders, endocrine metabolism, circadian rhythm disorders, cell growth regulation disorders, etc., through a search of the PubMed database. This search identified approximately 30 common disease types, most of which are related to environmental stress and adaptability. Using the OpenGWAS database and FinnGen^32^ (R9), the aforementioned diseases were further investigated, excluding those for which complete summary statistics could not be obtained. The study ultimately included 30 diseases (**Table 1**). Summary statistics from FinnGen and the UK Biobank, for diseases with available data from both datasets, were used for cross-validation. Diseases with a low number of SNPs and those lacking MAF in GWAS statistical data were excluded, resulting in 18 disease sets being included in the validation (**Table 1**).

### Mendelian randomization analysis

In this study, we employed a two-sample Mendelian randomization (MR) approach based on GWAS summary statistics to examine the causal relationship between plasma gene expression and the selected 30 diseases. For cases where a single SNP served as the genetic instrument (i.e., only one eQTL available for a given drug gene), the Wald ratio was used to estimate the causal effect. When two or more genetic instruments were available, inverse variance weighted MR (MR-IVW) was employed, assuming a zero intercept and conducting a weighted regression of the SNP-gene effects with the SNP-disease effects. Additionally, MR-Egger regression was utilized to test for horizontal pleiotropy between genetic instruments and diseases, and Cochran’s Q statistic was used to assess heterogeneity among the chosen genetic instruments. Both the MR regression beta and the odds ratio (log_10_(beta)) were obtained. Plasma gene expressions with p value<0.05 and consistent beta sign across the same disease were considered to have a causal relationship with the corresponding disease. Statistical analyses were conducted using R version 4.2.3, utilizing the TwoSampleMR packages.

### Reverse causation detection

To investigate potential reverse causality, plasma gene expressions identified as having a causal effect were included in bidirectional Mendelian randomization (MR) analyses. The analyses employed multiple methods to estimate the effects, including MR-IVW, MR-Egger, weighted median, simple mode, and weighted mode. Additionally, Steiger filtering was conducted to ensure the directionality of the association between gene expression and multiple diseases was correctly identified (*p* value < 0.05).

### Prediction of potential drugs for genes with causal effects

#### Drug prediction based on causal networks

Based on the causal gene network corresponding to each disease, two sets of opposing protein lists, one with beta greater than 0 and the other with beta less than 0, were utilized. Predictions for targetable drugs were made based on the SignatureSearch database, with criteria set at *p* value (up) < 0.01, *p* value (down) < 0.01, and Z score (sum) < −3. This method aims to explore a drug network that modulates the overall causal proteins in an antagonistic manner^33^. (**Table S1**)

#### Screening of drugs for clinical treatment of diseases

To explore targetable drugs for independent causal genes associated with diseases, the study identified a collection of 1,100 drugs used in clinical treatments from the Comparative Toxicogenomics Database (CTD), detailed in **Table S2**. Considering the potential adverse effects of non-clinical medications, the inclusion criteria for drugs were strictly controlled. Initially, drugs without targeted gene research were excluded, followed by those lacking a Chemical Abstracts Service Registry Number (CAS RN). Drugs were then selected based on the presence of ‘Direct Evidence’ marked as ‘T’ (True). Lastly, given the complexity of drug actions, only the top five drugs per disease, based on the number of references, were selected. This process ultimately included 53 drugs across 17 diseases, as shown in **Table S3**.

#### Determination of target genes for clinical therapeutic drugs

Data for 53 drug genes from the Comparative Toxicogenomics Database (CTD) has been accessed, focusing on their target proteins. Given eQTLs provide insights into gene expression, with GeneForms specified as mRNA, excluding instances of cotreatment, and the selection was narrowed to species including “Homo sapiens,” “Mus musculus,” “Peromyscus californicus,” “Rattus,” “Cricetulus griseus,” “Cavia porcellus,” “Cavia,” “Sus scrofa,” and “Rattus norvegicus.” Finally, to avoid drugs having a negative impact on diseases, our study only included drugs that, as the dosage increases, lead to changes in gene expression that have a positive effect on the disease (i.e., the relationship between the protein and the drug should be opposite to the relationship between the gene and the disease.

### Pathway enrichment studies

The study concentrated on drug genes and carried out Gene Ontology (GO) terms enrichment analysis via the Enrichment Map framework. This process entailed identifying significantly enriched pathways for individual disease sets (adjusted *p* value < 0.05) and utilized a clustering method with a CoSE Layout for categorizing and characterizing the pathways. The analyses were efficiently conducted leveraging the capabilities of Cytoscape’s AutoAnnotate and EnrichmentMap functions.

## Results

### Causal network of drug genes and multiple MDs

Leveraging known drug target genes, we aimed to explore effective causal genes across multiple diseases underscores the potential therapeutic targets for various conditions. Initially, our study utilized clinical research related to metabolic disorders, extracting GWAS summary data for 30 diseases from 377,277 individuals (210,870 females and 166,407 males) in the FinnGen database to perform Mendelian Randomization (MR, **Table 1**). We identified disease-related causal genes by examining blood eQTLs for 2,478 known drug target genes. Through sensitivity analysis (MR Egger) and consistency tests between IVW and Egger’s Q, along with verification of the causal effect direction (Steiger), MR incorporated 499 drug target genes with significant causal associations with 27 diseases (MR p value < 0.05, Q IVW and Q Egger >0.05, MR Egger> 0.05, Steiger *p* value< 0.05, |MR coefficient|> 0.2, **Fig. 1**). Among these, the *FN1* gene had the most disease connections, including 5 diseases (**Table 2**), where an increase in *FN1* protein was identified as a risk factor for Type 1 Diabetes (T1D)^34^, Polycystic Ovary Syndrome (PCOS)^35^, and a protective factor for Non-Alcoholic Fatty Liver Disease (NAFLD), Hypothyroidism/Myxedema, and Hyperthyroidism. The OBSCN gene was also linked to 5 diseases, with increased protein expression posing a risk for NAFLD, Colorectal Cancer^36^, Parkinson’s Disease, and serving as a protective factor for Preeclampsia and Non-Small Cell Lung Cancer^37^.

As shown in **Fig S1**, PCOS was associated with a greater number of causal genes, such as *CDKN1A*^38^, *TFRC*^39^, *MAPK8*^40^. Additionally, causal gene targets for T1D comprised *ICAM1*^41^, *NFKB1*^42^, and others. The study also uncovered potential causal targets for some cancers, for example, Small Cell Lung Cancer had 52 causal gene targets, including *VIM*^43^ and *TLR5*^44^. For psychiatric disorders, Schizophrenia included causal genes such as *NOS3*^45^, *ULK2*^46^, and *ALDH1A1*^47^ (**Table 2**). Using the UK Biobank (UKB) datasets, causal connections for only 10 diseases were verified, involving 31 genes (**Table S4**), due to the heterogeneity between the UKB and FinnGen populations.

### Functional clustering of key causal genes for multiple MDs

To explore the related functions of drug target genes with significant causal effects on diseases and the shared causal characteristics among diseases, our study conducted pathway function annotation for genes. 14 diseases demonstrated significant pathway enrichment (|MR coefficient| > 0.2, adjusted p value in pathway enrichment < 0.01), spanning 13 distinct pathway cluster categories. The causal genes for 13 diseases were annotated to pathways related to response stimulus regulation (**Fig. 1, Fig. S1B, Table S5**), including T1D, suggesting that intervening in the body’s stress response could be key to disease treatment. Uniquely in Alzheimer’s Disease (AD), our study focused on the ITGAV-VTN-ITGB5 pathway cluster, which includes various pathways related to the Integrin Subunit complex, such as ITGAV-ITGB5-VTN. The regulatory role of Integrin Subunit receptors is considered a key marker of aging. Additionally, the regulatory role of neurons was independently observed in AD. Lastly, NAFLD and AD were clustered into Positive peroxidase activity, highlighting potential therapeutic targets and the importance of oxidative stress pathways in these conditions.

### Exploration of clinical drugs for common key genes in multiple MDs

The regulation of drug target genes’ expression plays a pivotal role in the treatment of diseases. Our analysis revealed that key genes shared across multiple diseases may be regulated by a wider range of drugs (**Table 2** and **Table S6**), based on 53 clinical drugs. Within the response stimulus regulation pathway cluster, five genes were crucial for endocrine disorder diseases. The *FN1* gene, with causal connections to five diseases, was influenced by four drugs. Metformin, Troglitazone, and Zinc could potentially decrease *FN1* expression, offering therapeutic benefits for PCOS, while Genistein may increase *FN1* expression, offering a therapeutic effect for NAFLD. Additionally, the *ICAM1* gene was modulated by six drugs, showing therapeutic effects for Anxiety and NAFLD. The *EDN1* gene, also regulated by six drugs, presented therapeutic potential for Parkinson’s Disease, Anxiety, and Non-Small Cell Lung Cancer. Furthermore, the *CDKN1A* gene was affected by five drugs, indicating therapeutic effectiveness for PCOS. Lastly, significant causal associations with two nervous system diseases were observed with the Vimentin (*VIM*) gene, whose expression is co-downregulated by Sirolimus and Estradiol. This comprehensive analysis underscored the complex interplay between drug action and gene expression regulation, highlighting the potential for repurposing drugs across a spectrum of diseases by targeting key regulatory genes.

### Discovery of potential therapeutic mechanisms of MD clinical drugs

Diseases can be influenced by the expression of multiple genes and simultaneously modulated by various drugs. Hence, exploring the targetable drug-gene causal network, we investigated the KEGG pathway functions of drug target genes. Among 53 clinical drugs for multiple diseases, we identified 230 ‘drug - gene expression - disease’ connections (**Table 3, Fig. S7**). Valproic Acid regulated the expression of 20 genes and had a causal connection with Schizophrenia, where *SLC29A1* and *HDAC4* were enriched in Alcoholism, potentially involved in neuroexcitation regulation. Doxorubicin’s modulation of 13 gene expressions was causally linked to Colorectal Cancer, with *IL7R*, *STAT3*, *FLT1* enriched in HIF-1 signaling pathway and FoxO signaling pathway, involved in the activation regulation of cellular HIF factors. Additionally, NAFLD had a causal effect with 12 genes, enriched in the AGE-RAGE signaling pathway in diabetic complications (adjusted p value < 0.05), covering three genes, with Genistein potentially increasing *FN1* and *NOS3* expression to prevent liver fibrosis and cirrhosis. In PCOS, 12 genes had causal links with drugs, where *FN1*, *ICAM1*, *NOS3* were modulated by Ethinyl Estradiol’s gene expression, related to Legionellosis, and possibly involved in pro-inflammatory response and cell chemotaxis post bacterial infection. Moreover, genes like *MAPK8*, *CASP3*, *CXCL1*, *HSP90AA1*, regulated by Zinc, were associated with the progression of PCOS and participate in IL-17 signaling. Lastly, we observed associations among eight genes in Parkinson’s Disease, where *ITGAM* and *HLA-DRB1* significantly indicated the activation process of LPS, related to inflammatory responses. This comprehensive analysis highlighted the intricate mechanisms through which drugs modulated gene expression and their potential therapeutic effects on various diseases through specific pathway involvements.

### Disease-based causal proteins discover new uses for old drugs

Based on the influence of drugs on gene expression and the significant causal association between gene expression and disease occurrence (|MR coefficient| > 0.2), our findings suggested strategies for repurposing existing drugs for new therapeutic uses. The study identified 1,111 new ‘Drug-Gene-Disease’ causal connections (**Table S8**). Notably, drug repurposing opportunities were observed in four diseases: Bipolar Disorder, Manic Episode, Small Cell Lung Cancer, and T1D. Specifically, 41 genes were found to have a causal effect on T1D, with KEGG functional enrichment (**Table S9**) indicating potential new therapeutic pathways, including the PI3K-Akt signaling pathway and ECM-receptor interaction (**Table S10, Fig. S2 A(b) (c)**). Nine drugs were involved in regulating genes within the PI3K-Akt signaling pathway, with Aspirin recently reported as a protective drug for diabetes^48^ (**Fig. S2 A(a)**).

Currently, there are no targeted drugs for Manic Episode, but our research suggested 61 genes have a causal effect on Manic Episode. Functional enrichment also indicated that ECM-receptor interaction and Kaposi sarcoma-associated herpesvirus infection could be potential new therapeutic pathways, involving genes such as *COL6A1*, *COL6A2*, *VWF*, *THBS3*, *CCR1*, *EP300*, *HLA-G*, *PLCG2*, *FAS*, *PDGFB*, with 28 drugs modulating the Kaposi sarcoma-associated herpesvirus infection pathway (**Fig. S2 B(b) (c)**). Notably, based on the shared pathway of ECM-receptor interaction between Manic Episode and T1D, four common new drugs were identified: Cannabidiol, Doxorubicin, Genistein, Propylthiouracil.

Furthermore, predictive analyses based on existing drugs were conducted across 499 causal genes associated with metabolic disorders in different diseases, leading to a comprehensive drug prediction targeting these genes. We identified 189 drugs (p (up) < 0.01, p (down) < 0.01, Z score (sum) < −3, **Table S1**). Our results highlighted the potential collective modulation effects of some drugs on causal gene sets across multiple diseases (**Fig. S1 C**), with Reserpine showing potential therapeutic capabilities for Alzheimer’s Disease and NAFLD. Additionally, Bortezomib might have potential therapeutic effects on PLOS, NAFLD, and Schizophrenia, suggesting a broad spectrum of action and the possibility of drug repurposing for complex disorders.

## Discussion

To our knowledge, this study represents the pioneering effort to integrate eQTL analysis for uncovering pivotal drug-gene causal relationships in metabolic disorders, employing two-sample Mendelian Randomization (MR) alongside drug prediction techniques. Herein, we identified multiple potential drug target genes with causal associations to metabolic disorders, such as *FN1*, *ICAM1*, *SNCA*, *SLC29A1*, and *VIM*. The causal associations of some genes with diseases were validated in the UK Biobank (UKB) dataset, including *ICAM1* and *SLC29A1*. Additionally, we provided potential therapeutic mechanisms for various diseases through clinical drugs, such as the treatment of Schizophrenia with Valproic Acid, potentially based on the neuroexcitation regulation by *SLC29A1* and *HDAC4*. Genistein may increase the expression of *FN1* and *NOS3* to prevent liver fibrosis and cirrhosis, offering a treatment for NAFLD. Lastly, based on the MR causal network, we proposed drug repurposing strategies for diseases, focusing on four conditions. Manic Episode and T1D share common drug action pathways and four new common drugs, highlighting the potential for cross-disease therapeutic applications.

This study highlights several key genes with robust associations across metabolic disorder-related diseases, suggesting *FN1* (fibronectin 1), *ICAM1* (Intercellular Adhesion Molecule 1), and SNCA (Synuclein Alpha) as potential critical targets for these conditions. Research indicates *FN1* as an adverse marker for PCOS^35,49^ and T1D^34^, consistent with the observed increased prevalence of metabolic features (diabetes, hypertension, etc.) in women with PCOS^50–52^. Our findings suggested a negative correlation between *FN1* and NAFLD, where upregulation of *FN1* expression may suppress excessive inflammation and repair liver function in many NAFLD patients. Some studies have hinted that thyroid hormones could increase fibronectin 1 expression through the PI3K/Akt/HIF-1 pathway^53^, though research on its feedback effects on thyroid function is yet to be reported. Our results also show a strong causal effect of *ICAM1* with T1D, T1DW, and NAFLD. Located within a diabetes linkage region^41^, *ICAM1* primarily participates in integrin receptor activation. Increased expression of the *ICAM1* gene has been associated with all-cause mortality and cardiovascular disease incidence in T1D patients^54,55^. Moreover, an elevated concentration of *ICAM1* protein serves as a significant diagnostic marker for NAFLD^56^. Although the protective effect of *ICAM1* on Anxiety is a novel possible causality, it has been validated by analysis using UK Biobank data, with no previous studies reported. Furthermore, our analysis suggested that SNCA offers protection against PCOS but may pose a risk factor for AD. While some studies have reported correlations between PCOS and AD^50^, definitive answers remain elusive. Increases in SNCA protein have been suggested as molecular markers for AD^57,58^, while positively correlating with protective HSP90 proteins in PCOS patients^59^. Additionally, we found opposing causal relationships of *APP* (Amyloid Beta Precursor Protein) with AD and PCOS. Lastly, the protective role of *INSR* for AD^60^ and its risk association with Gestational Diabetes (GD)^61^ have been confirmed, with UK Biobank validating the association of *INSR* with Hypothyroidism. We also noted a significant association between *ITGAM* (CR3) and Parkinson’s Disease (PD)^62^, adding to the understanding of the genetic underpinnings and potential therapeutic targets across these metabolic and neurological disorders.

Our pathway analysis extensively focused on response stimulus regulation. Previous literature indicated that metabolic disorders are associated with resistance to stimuli, both external^63^ and internal^29^. Meta-analyses suggested possible concurrent mechanisms among some of these diseases^64–67^, leading us to believe that metabolic dysregulation induced by response stimulus regulation is significant evidence of the association among metabolic disorders. Additionally, in the context of Alzheimer’s Disease (AD), significant enrichment was found in the PTD domain neuronal, where APP and INSR were identified as the main causal genes, consistent with previous research^68,69^. Furthermore, in AD, we discovered evidence related to the itgav vtn itgb cluster (*ITGAV*, *ITGB5*, *LTBP2*), though such associations have not been reported in existing literature. However, the role of integrin receptors in AD has been confirmed^70^, suggesting our findings may point to novel therapeutic pathways for AD treatment. This underscores the importance of investigating the complex interplay between genetic factors and metabolic processes, paving the way for innovative approaches to understanding and treating metabolic disorders and related diseases.

Based on the constructed causal network for metabolic disorders, we explored the potential therapeutic mechanisms of known clinical drugs for each disease. The reduction in *SLC29A1* (Equilibrative nucleoside transporter type 1) expression, which can affect glutamate transport^71^, has been causally associated with Schizophrenia, a finding validated in the analysis using UK Biobank datasets. Current research suggests that the effect of Valproic Acid on Schizophrenia also involves modulation of glutamate transport, with our results indicating a novel pathway involving Acute ethanol through the *SLC29A1* and *HDAC4* genes. Moreover, the effect of Doxorubicin on Colorectal Cancer has been extensively reported^72^. We discovered that Doxorubicin’s reduction of *STAT3* and *FLT1* expression within the HIF-1 signaling pathway might effectively inhibit the activation of *HIF1A* and angiogenesis, potentially representing an important therapeutic approach for Colorectal Cancer (**Fig. S3 B**). This aligns with research suggesting Doxorubicin’s inhibition of HIF as an effective treatment for breast cancer^73^, consistent with our findings. Additionally, studies showed Genistein treats NAFLD by targeting Thromboxane A2. Our findings regarding its effect on the expression regulation of genes related to Apoptosis (*NOS3*), Thrombogertesis inflammation atherosclerosis^74^ (*ICAM1*^56^), and Mesangial matrix expansion (*FN1*) may suggest new drug treatment mechanisms for NAFLD (**Fig. S3 C**). We also noted that Copper might modulate the phase in Circadian entrainment through downregulation of *PRKG2* and *CALM3*, indicating a possible treatment pathway for Colorectal Cancer. Consistently, research suggested that disruptions in circadian rhythms could affect the likelihood of Colorectal Cancer^75,76^ (**Fig. S3 D**), highlighting the complex interplay between genetic expressions, drug effects, and disease outcomes.

Drug repurposing is considered a safer approach because it leverages existing knowledge about drug dosage, safety, and side effects, significantly enhancing the efficacy of clinical trials^77^. Exploring drug repurposing based on drug databases and clinical drugs used in metabolic disorders, we identified potential new uses for existing drugs in Manic Episode and T1D. The mechanism of action for drug repurposing in both conditions involves a common pathway: ECM-receptor interaction. Research indicated that the interaction between ECM and receptors can alleviate diabetic nephropathy^78^, and cell therapy for T1D also underscores the importance of ECM^79^. For T1D, the gene expression in this pathway could be potentially modulated by six drugs, with Cannabidiol, Doxorubicin, Genistein, and Propylthiouracil being of interest in the same pathway for Manic Episode. Clinical trials have suggested the potential adjunctive therapeutic effects of Cannabidiol for Manic Episode^80^ and T1D^81^. Besides, Genistein has been reported as a potential alleviator for both T1D and T2D^82^. Additionally, it is noteworthy that we have identified a potential association between the recently disclosed diabetes^83^ therapeutic drug and T1D. However, the mechanisms of action for these drugs have yet to be fully elucidated. Leveraging the existing clinical drugs combined with the causal network of multiple diseases, we identified potential therapeutic drugs for Manic Episode and T1D and substantiated the feasibility of these drugs based on literature evidence. Our research proposed the potential for repurposing existing drugs to provide new therapeutic options for complex diseases, thereby accelerating the path to effective treatments.

Inevitably, our study comes with its set of limitations, including the constrained validation of diseases within the UK Biobank and an emphasis on European populations. Such specificity may lead to a variation in findings across different ethnic groups, a reflection of the vast spectrum of genetic diversity. Moreover, while our strategy for drug repurposing in metabolic disorders is informed by initial insights, the comprehensive molecular mechanisms and the safety profiles of the proposed medications demand further in-depth investigation. Looking forward, we are optimistic that our work could help pave the way for more inclusive and holistic studies in the future, ultimately leading to the identification of treatments with universal efficacy.

## Data Availability

All data generated are available online

https://quantlifebits.shinyapps.io/mrmetabopharm/

## Ethics approval and consent to participate

Not applicable.

## Consent for publication

All authors agree to publish.

## Availability of data and materials

We publish all the causal network and drug network information in the form of website on: https://quantlifebits.shinyapps.io/mrmetabopharm/.

## Authors’ contributions

SZ, XH designed most of the studies. DS, XS and DH carried out much of the work together. SZ, XH and XS analyzed the data. SZ and DH provided revision of the figure. XH and XS complete methodology-related content. DS provided insights into the discussion section. All authors wrote up the manuscript and approved the final manuscript.

## Acknowledgements

Thanks to FinnGen and UK Biobank for providing data support.

---

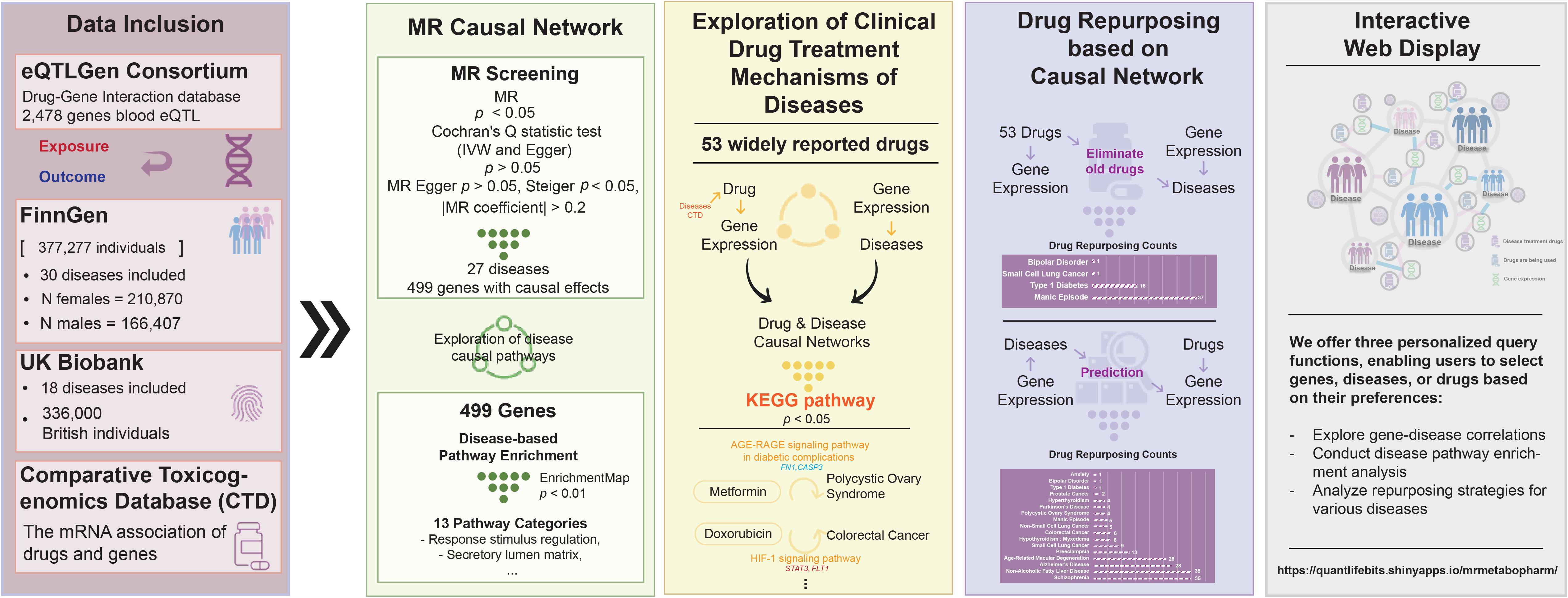

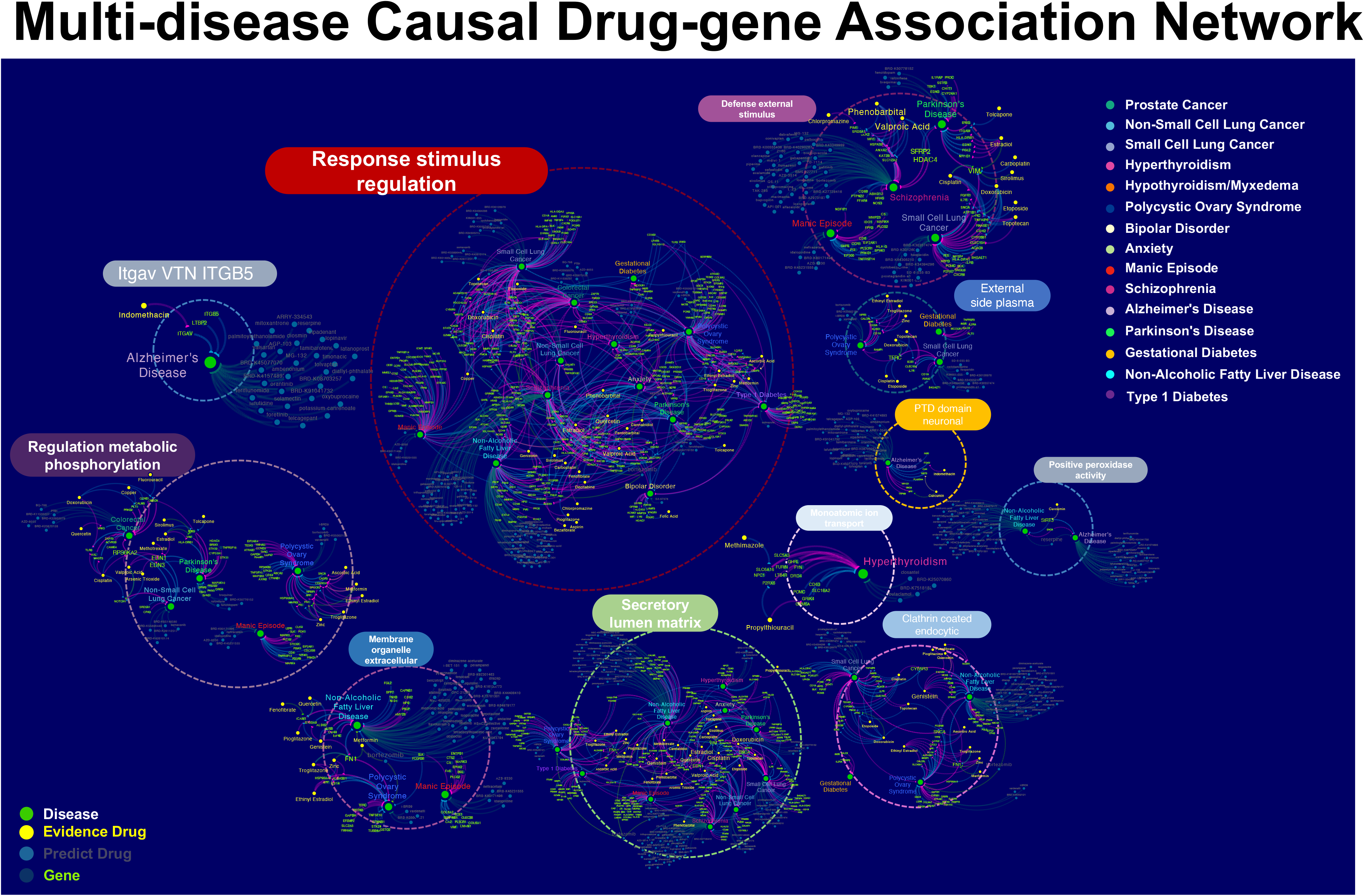

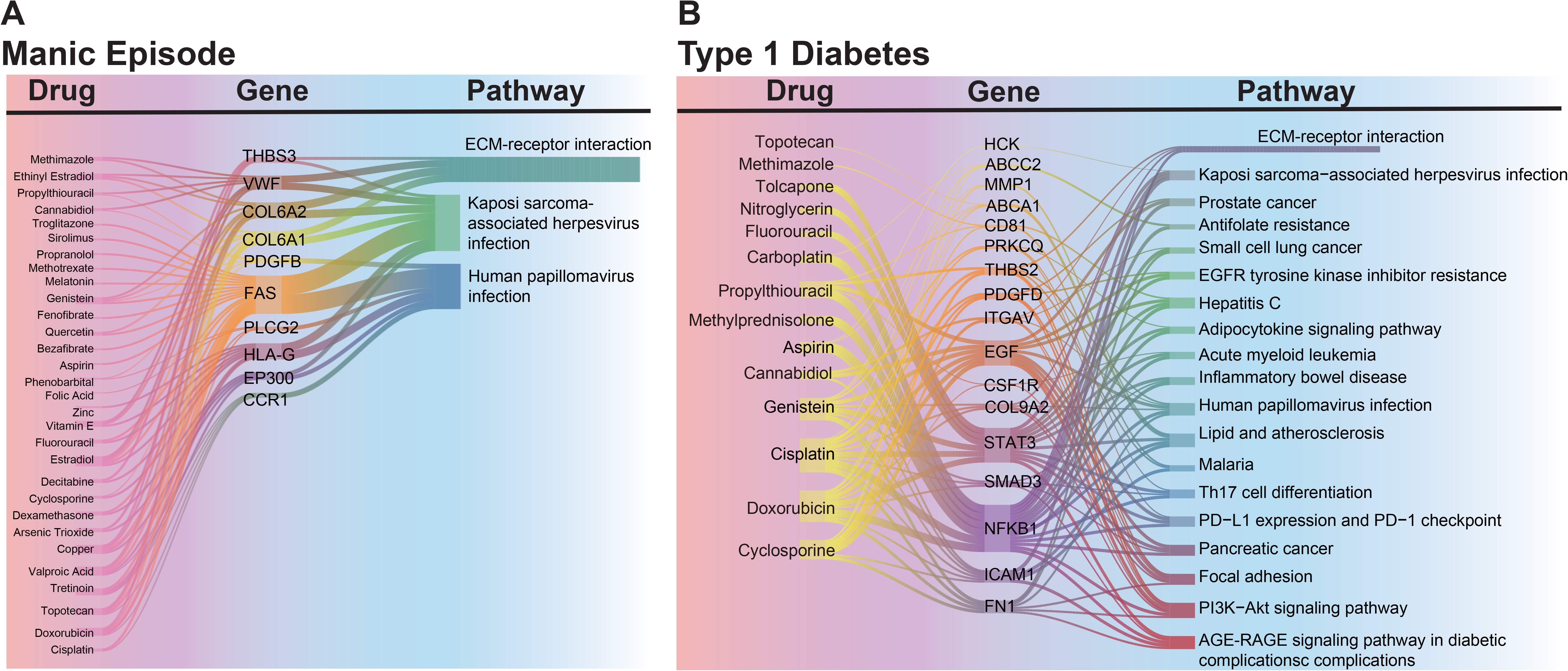

