## Supplementary material for "Pharmaceutical Repurposing Strategies for Metabolic Disorders: Insights from Mendelian Randomization Studies": FigS1

# A
### Number of causal genes for Disease MR

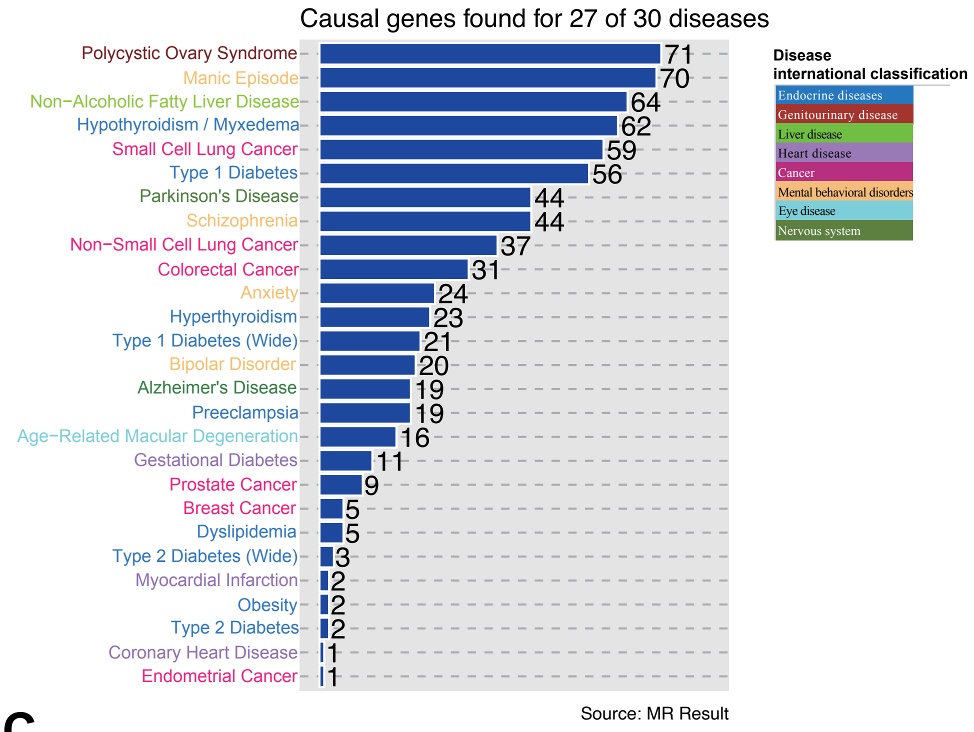


# B
### Functional enrichment analysis of Disease causal association genes

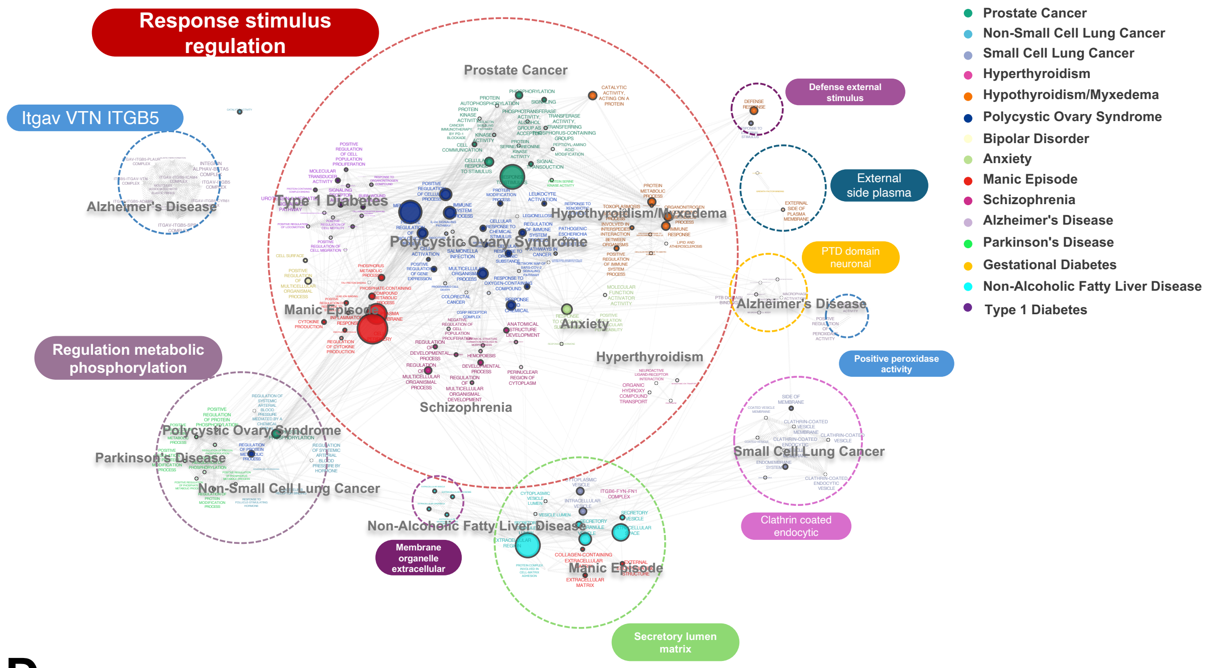


# C
### Disease Drug Prediction Network

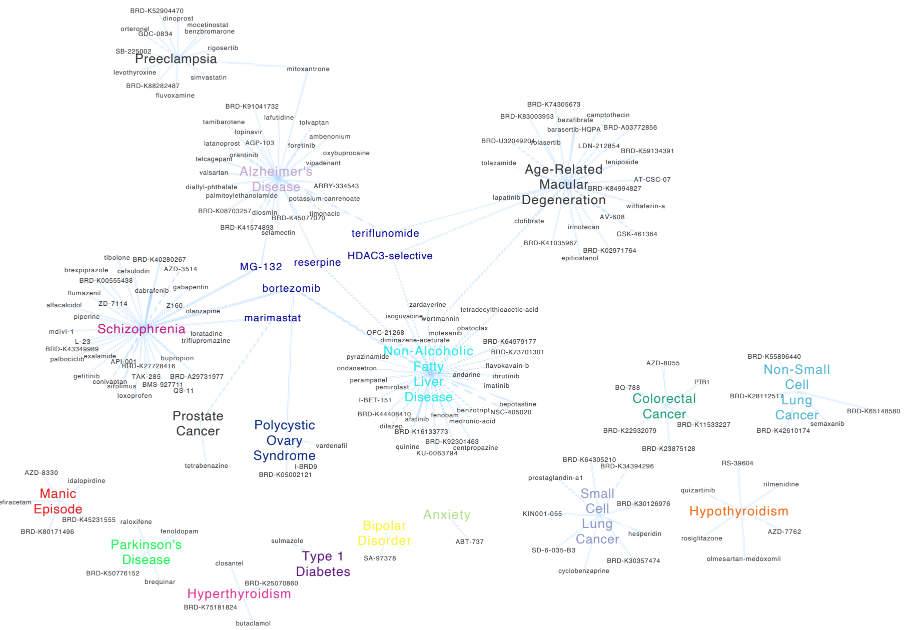


# D
### Disease drug prediction frequency distribution

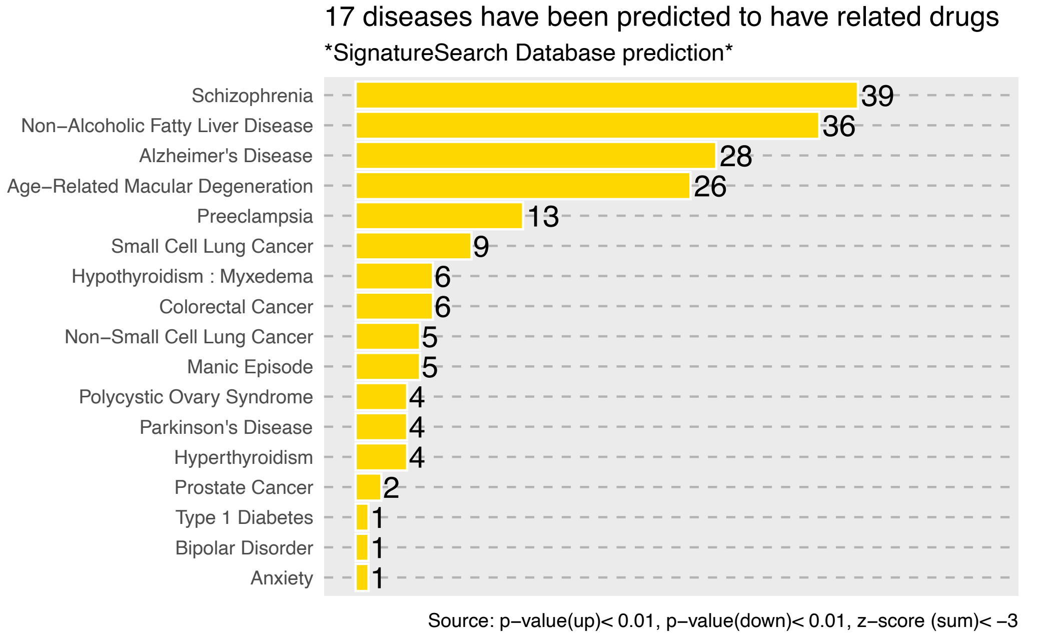
