## Supplementary material for "Pharmaceutical Repurposing Strategies for Metabolic Disorders: Insights from Mendelian Randomization Studies": FigS2

**A**

(a) Type 1 Diabetes Old medicines repurposed

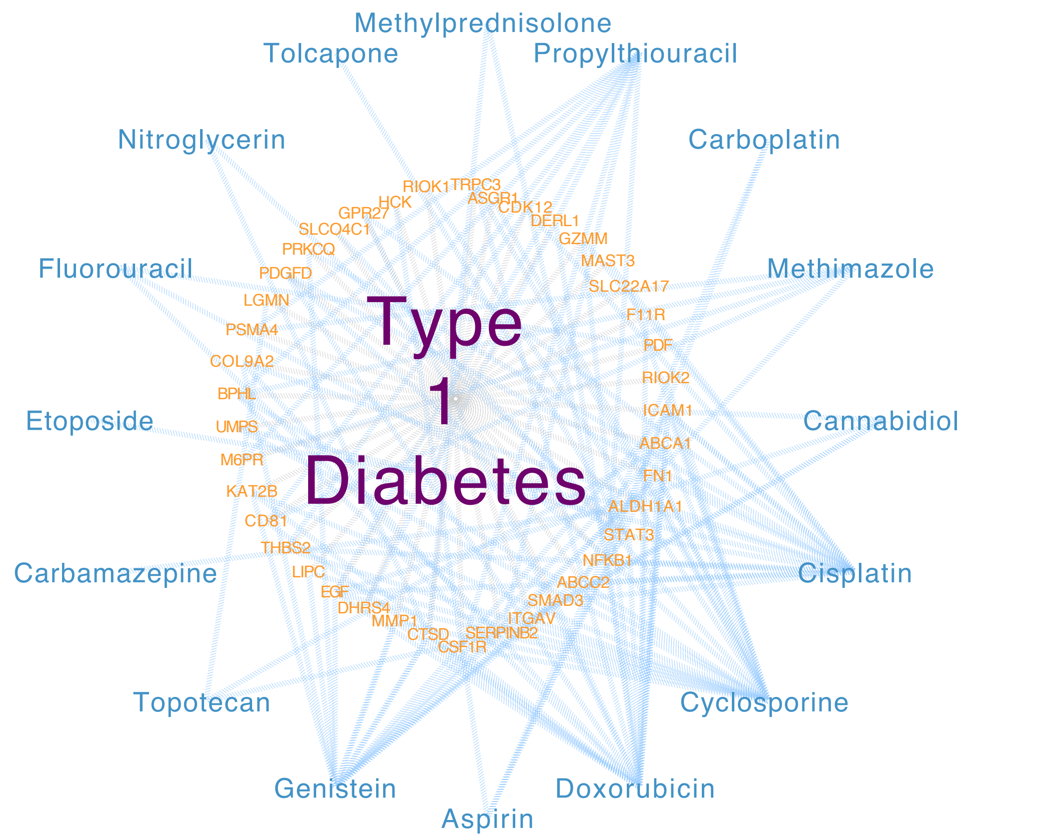


(b) KEGG in PI3K-Akt signaling pathway

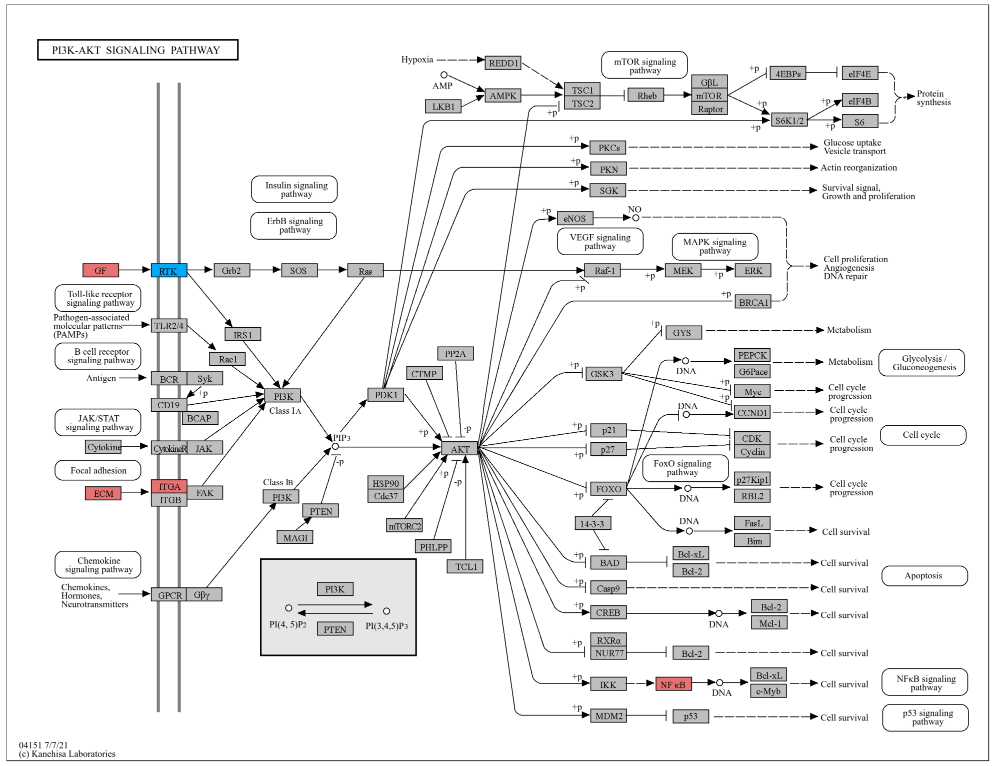


**Cisplatin, Cyclosporine, Cannabidiol, Doxorubicin, Propylthiouracil, Methylprednisolone, Genistein, Carboplatin, Aspirin**

(c) KEGG in ECM-receptor interaction

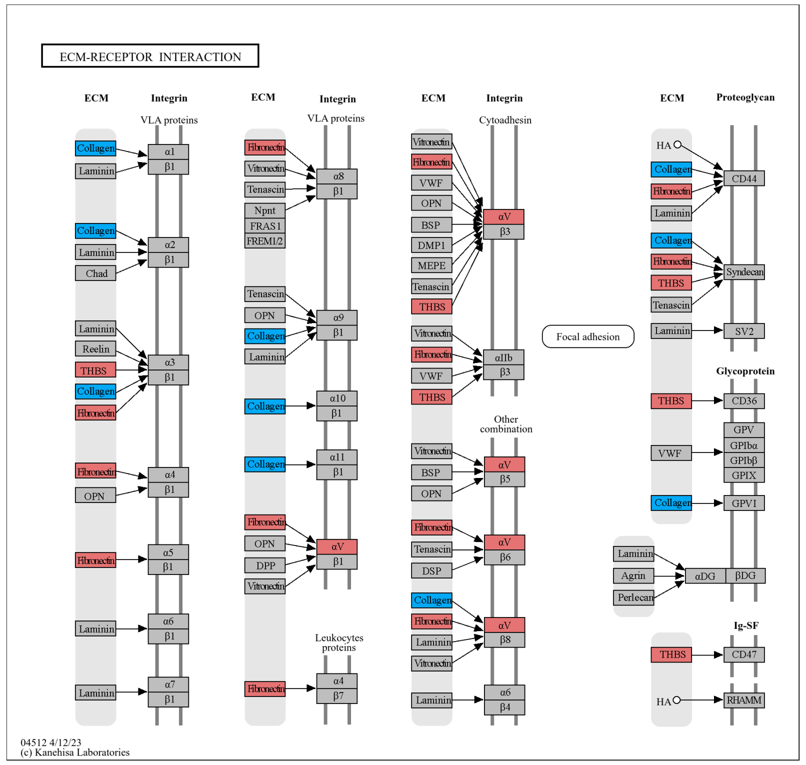


**Cisplatin, Cyclosporine, Cannabidiol, Doxorubicin, Genistein, Propylthiouracil**

**B**

(a) Manic Episode Old medicines repurposed

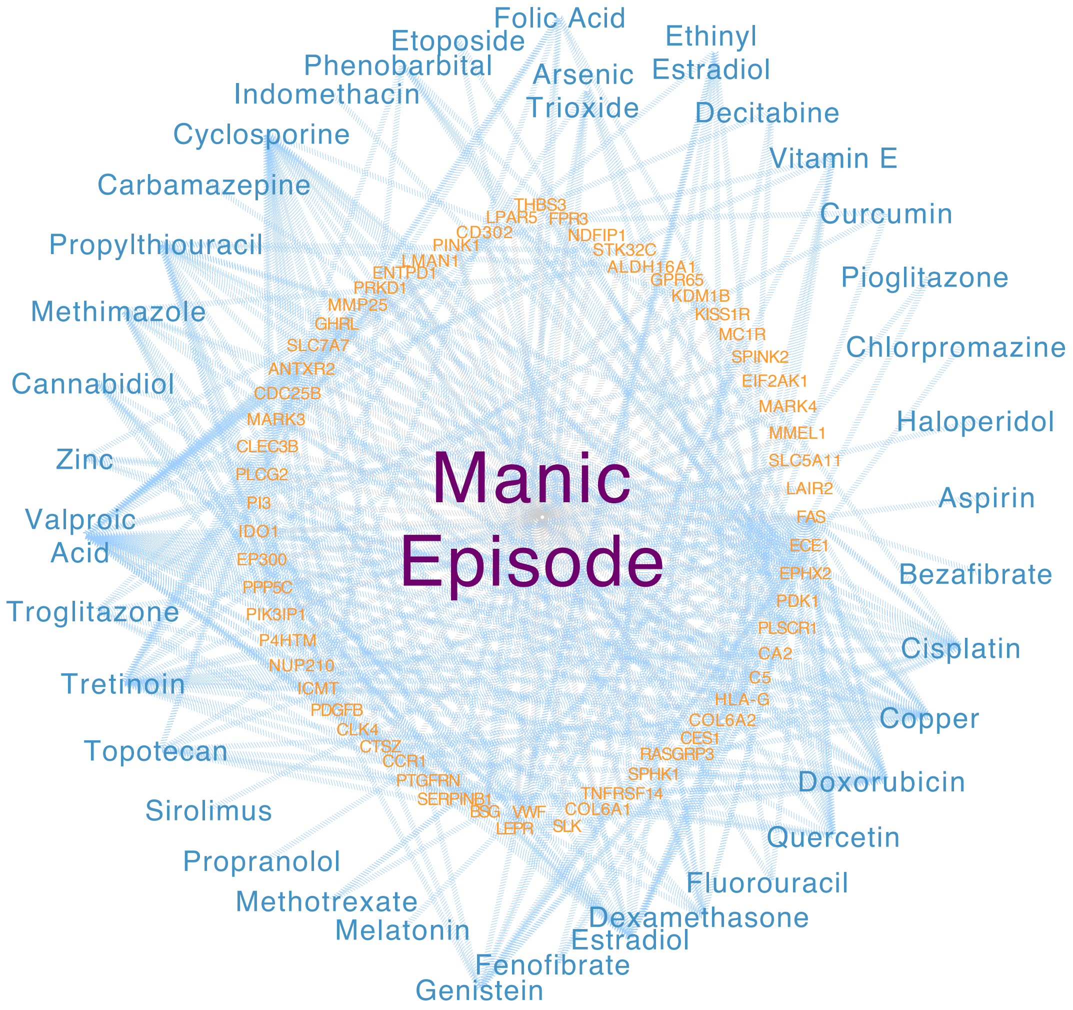


(b) KEGG in Kaposi sarcoma-associated herpesvirus infection

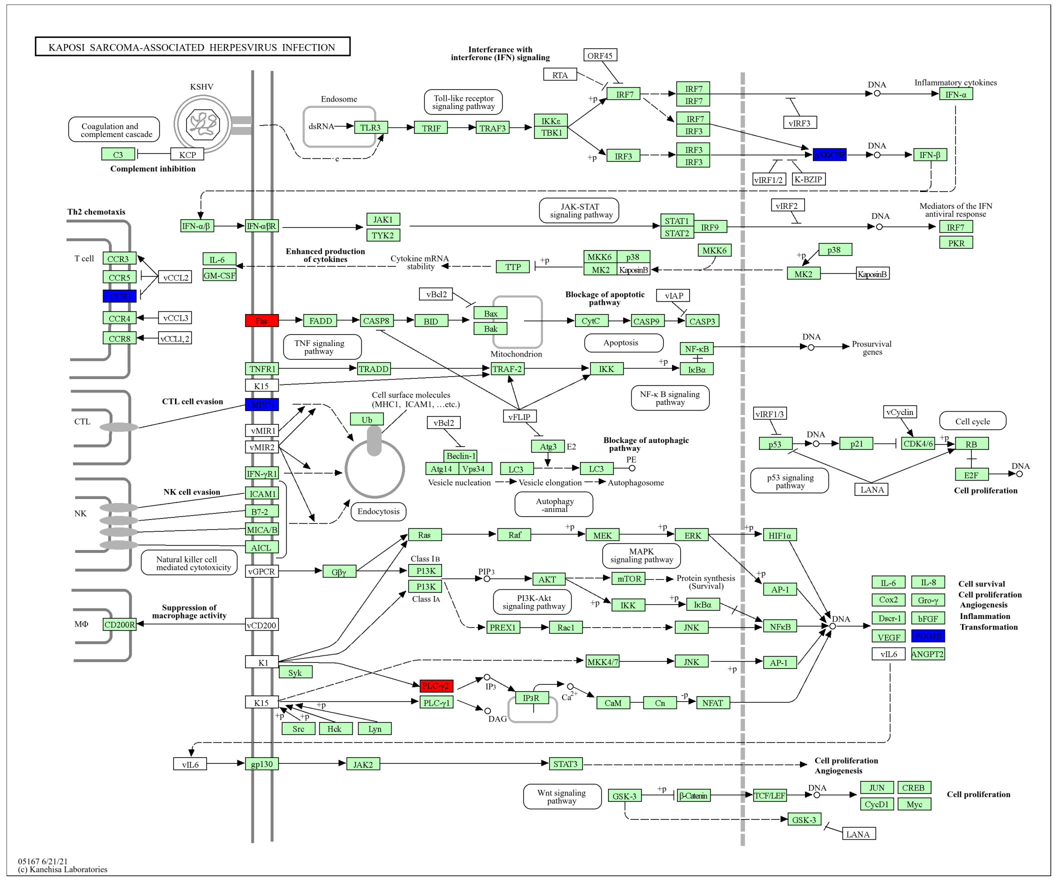


**Cisplatin, Copper, Cyclosporine, Aspirin, Cannabidiol, Doxorubicin, Arsenic Trioxide, Decitabine, Bezafibrate, Topotecan, Dexamethasone, Estradiol, Folic Acid, Genistein, Tretinoin, Fluorouracil, Phenobarbital, Valproic Acid, Propylthiouracil, Quercetin, Vitamin E, Zinc, Fenofibrate, Melatonin, Methotrexate, Propranolol, Sirolimus, Troglitazone**

(c) KEGG in ECM-receptor interaction

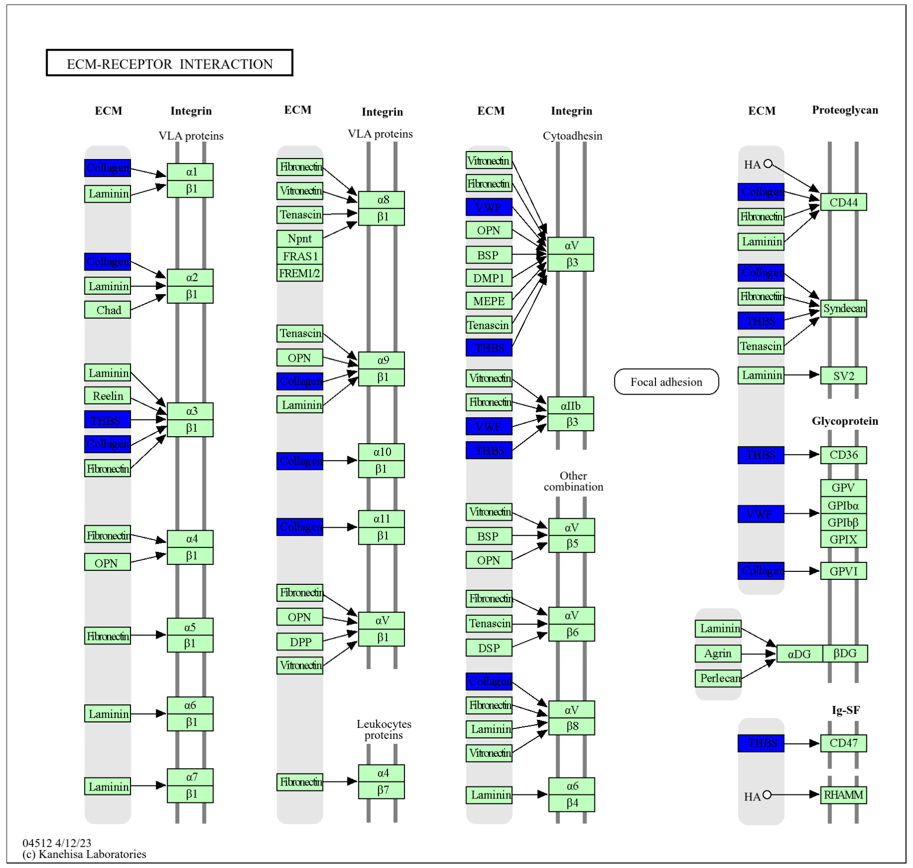


**Doxorubicin, Copper, Arsenic Trioxide, Estradiol, Cannabidiol, Decitabine, Ethinyl Estradiol, Genistein, Quercetin, Methimazole, Topotecan, Propylthiouracil, Valproic Acid, Vitamin E**
