## Supplementary material for "Pharmaceutical Repurposing Strategies for Metabolic Disorders: Insights from Mendelian Randomization Studies": FigS3

**A**

Alcoholism for Valproic Acid treatment Schizophrenia

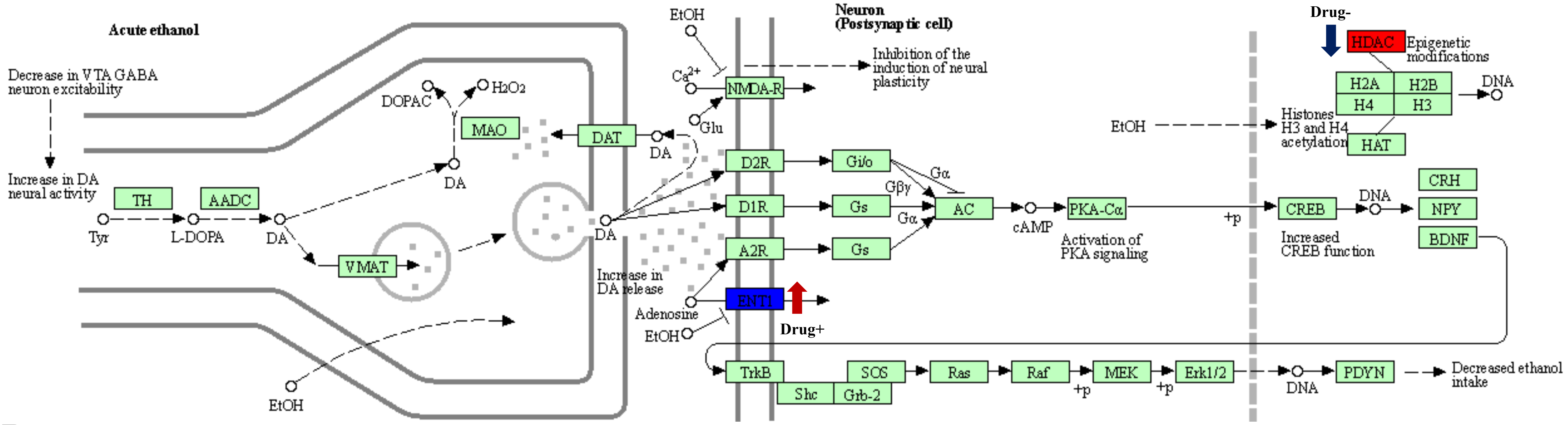


**B**

HIF-1 signaling pathway for Doxorubicin treatment Colorectal Cancer

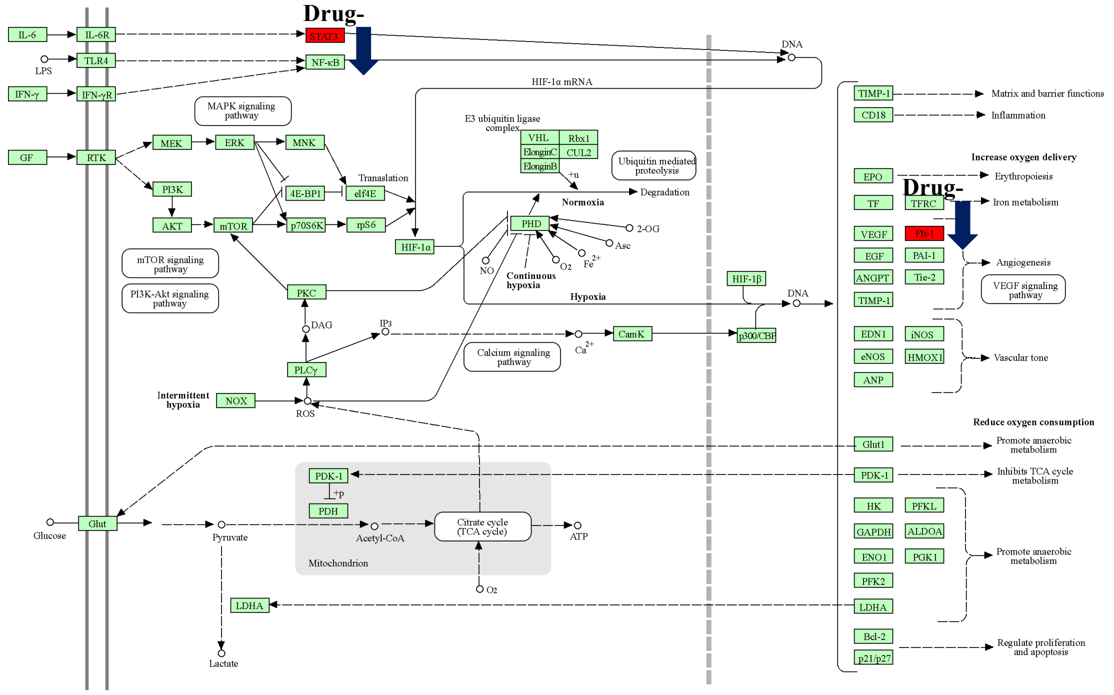


**C**

AGE-RAGE signaling pathway in diabetic complications for Genistein treatment NAFLD

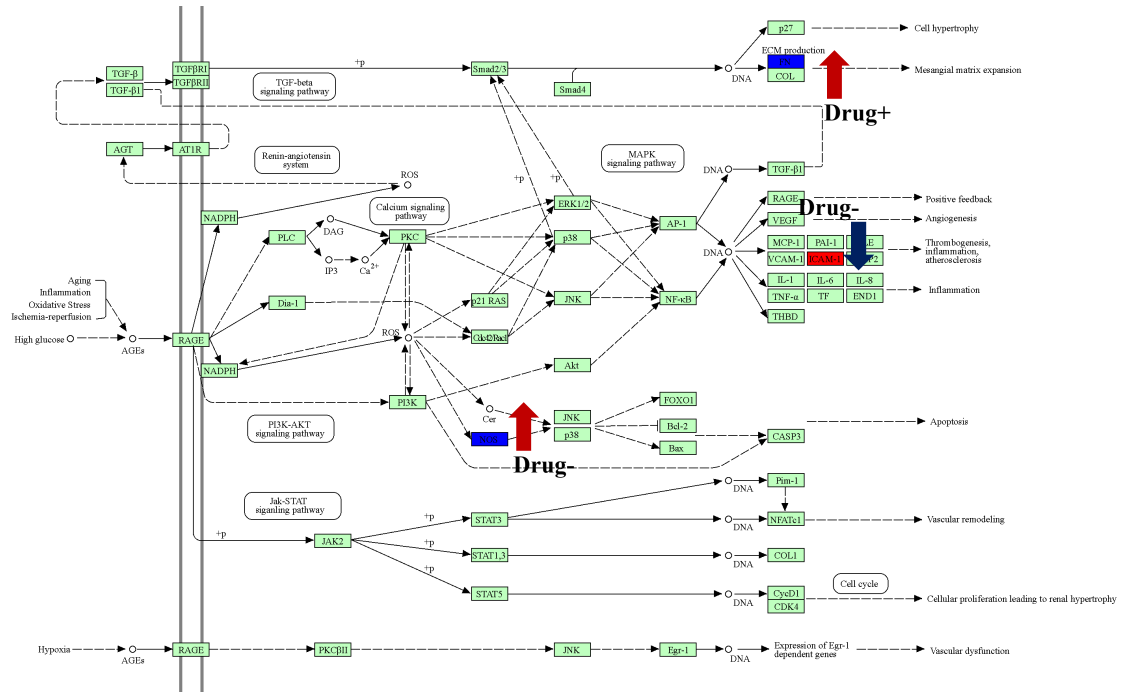


**D**

Circadian entrainment for Copper treatment Colorectal Cancer

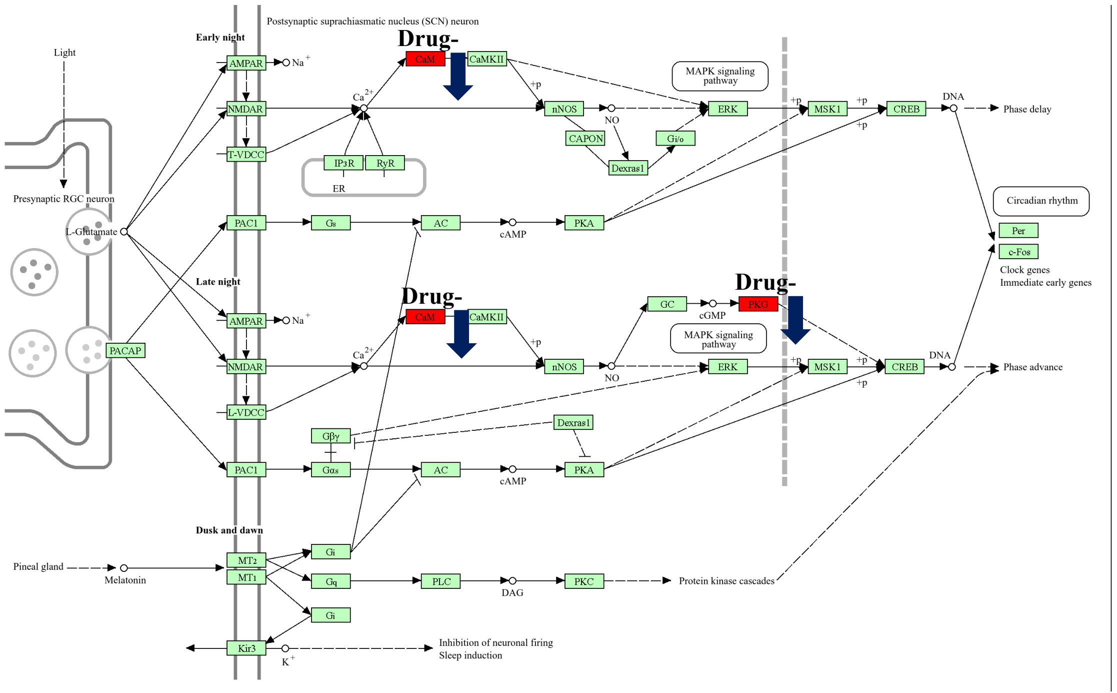
